# Prevalence and predictors of microvascular and macrovascular diabetes complications in adult Ugandans: a systematic review and meta-analysis

**DOI:** 10.1101/2024.10.15.24315542

**Authors:** Davis Kibirige, Ronald Olum, William Turyamureeba, Bethan Morgan, Andrew Peter Kyazze, Yakobo Nsubuga, Jerom Okot, William Lumu, Felix Bongomin

## Abstract

**Introduction:** There is a growing prevalence of diabetes and related chronic complications in Uganda. We conducted a systematic review and meta-analysis to document the prevalence and predictors of five microvascular and macrovascular diabetes complications in adult Ugandans with diabetes.

**Materials and Methods:** We searched Medline, EMBASE, CINAHL, Cochrane Library, and Africa Journal Online databases. We included studies on the prevalence and predictors of any chronic microvascular or macrovascular diabetes complications of interest. We conducted a random effect meta-analysis to determine the pooled prevalence of each diabetes complication. A narrative review was used to describe the significant predictors.

**Results:** A total of 20 studies involving 11,400 participants were included. The pooled mean (standard deviation) age of the participants was 54.8 (3.6) years, with the majority being female (pooled proportion of 61.1%, 95% confidence interval [CI] 57.1-65.2). For the microvascular diabetes complications, the pooled prevalence of diabetic neuropathy, retinopathy, and nephropathy was 56.8% (95% CI 44.9-68.7, I^2^ = 98.56%, p<0.001), 19.5% (95% CI 3.9-35.2, I^2^ = 99.60%, p<0.001), and 17.7% (95% CI 7.3-28.0, I^2^ = 99.36%, p<0.001), respectively. For the macrovascular diabetes complications, the pooled prevalence of peripheral arterial disease and diabetic foot disease was 32.2% (95% CI 15.8-48.7, I^2^ = 97.67%, p<0.001) and 5.5% (95% CI 1.7-9.2, I^2^= 90.22%, p<0.001), respectively.

Hypertension comorbidity, physical inactivity, family history of diabetes, body mass index ≤30 kg/m^2^, and pregnancy were predictors of diabetic nephropathy in three studies. In two studies, a history of a foot ulcer and age >60 years were predictors of diabetic neuropathy while female sex, hypertension comorbidity, and use of glibenclamide were predictors of peripheral arterial disease.

**Discussion:** Chronic diabetes complications are very common in adult Ugandans with diabetes, especially diabetic neuropathy and peripheral arterial disease. Regular screening and optimal management of diabetes and its complications should be emphasised in Uganda.

## INTRODUCTION

Globally, there is an increasing burden of diabetes. According to the most recent International Diabetes Federation (IDF) estimates, about 24 million (1 in 22) adult Africans aged between 20 and 79 years live with diabetes. Additionally, among the IDF regions, Africa has the highest proportion of people with undiagnosed diabetes (54%) [1]. Because of the high rates of undiagnosed diabetes in Africa coupled with low access to essential diabetes medicines and diagnostics and poorly structured healthcare systems, the prevalence of diabetes-related complications in adult Africans with diabetes is considerably high [2, 3].

Uganda has documented a two-fold increase in the prevalence of diabetes and prediabetes over the last ten years [4, 5]. Currently, the proportion of adult Ugandans who are unaware of their diabetes status is still relatively high [5]. Furthermore, access to affordable essential diabetes medicines and diagnostics remains challenging [6–8]. Because of these factors, the prevalence of chronic diabetes complications in adult Ugandans with diabetes continues to increase steadily.

Several observational studies have been conducted in Uganda to document these chronic diabetes complications, with significant variations in reported prevalence. In this study, we conducted a comprehensive systematic review and meta-analysis to document the pooled prevalence and predictors of chronic diabetes complications in adult Ugandans with diabetes to inform clinical practice and policy interventions on routine screening and management of diabetes in Uganda.

## MATERIALS AND METHODS

This systematic review and meta-analysis was conducted according to the criteria outlined in the Preferred Reporting Items for Systematic Reviews and Meta-Analyses (PRISMA) statement [9]. The PRISMA checklist is available as a supplementary Table 1. The study protocol was registered in the PROSPERO International Prospective Register of Systematic Reviews (CRD42024541251).

### Search strategy

With the help of a librarian (BM), we searched PubMed, EMBASE, CINAHL, Cochrane Library, and Africa Journal Online databases for published studies from 1946 to 2^nd^ May 2024. The following search terms were used “Diabetes mellitus’’ OR ‘’diabetes mellitus type 2’’ OR “diabetes” OR “diabetes mellitus type 1” OR “diabetes mellitus type 2” OR “type 2 diabetes” OR “type 2 diabetes mellitus” OR “type 1 diabetes” OR “type 1 diabetes” OR “type 1 diabetes mellitus” OR “diabetic” OR “type 2 diabetic” OR “type 1 diabetic” AND “microvascular complication” OR “macrovascular complication” OR “microvascular diabetic complication” OR “macrovascular diabetic complication” OR “microvascular diabetes complication” OR “microvascular diabetes complication” OR “microvascular” OR “macrovascular” OR “chronic diabetes complication” OR “diabetic retinopathy” OR “retinopathy” OR “maculopathy” OR “diabetic nephropathy” OR “nephropathy” OR “microalbuminuria” OR “macroalbuminuria” OR “albuminuria” OR “diabetic kidney disease” OR “diabetic neuropathy” OR “neuropathy” OR “sensory neuropathy” OR “peripheral neuropathy” OR “diabetic peripheral neuropathy” OR “diabetic sensorineuropathy” OR “peripheral arterial disease” OR “atherosclerotic vascular disease” OR “diabetic foot” OR “diabetic foot ulcer” OR “foot ulcers” AND “Uganda”.

In addition to the search of the databases above, we also hand-checked the references of the articles whose full texts were retrieved for any additional studies.

### Study selection criteria

The preliminary screening of titles and abstracts to identify potentially eligible articles was done independently by two reviewers (YN and FB) after removing duplicates. In case of any disagreement, an independent reviewer (RO) was consulted.

Full texts of the potentially eligible studies were retrieved and reviewed for the information of interest by three reviewers (DK, WT, and WL). The inclusion criteria of studies were: randomised controlled trials, cohort, case-control, cross-sectional, and retrospective studies published between 1946 to 2^nd^ May 2024 in the English language with any information on the sociodemographic (age, sex, family history of diabetes, and history of current smoking), clinical (duration of diabetes, co-existing HIV, and hypertension, glucose-lowering therapies being used, systolic blood pressure [SBP] and diastolic blood pressure [DBP]), anthropometric (body mass index or BMI), and metabolic (fasting blood glucose [FBG] and/or glycated haemoglobin [HbA1c]) characteristics, prevalence, and predictors of the chronic diabetes complications of interest.

### Data extraction

After identifying the eligible studies, the relevant study information of interest was extracted by one author (DK) using a Microsoft Excel 2016 form. The information that was extracted included: the last name of the first author and year of publication, the number of study participants, the number and proportion of female participants, the proportion of participants with a family history of diabetes, current history of smoking, self-reported co-existing hypertension, and HIV. We also extracted information on the glucose-lowering treatment being used (metformin, metformin-sulfonylurea combination, and insulin therapy either as monotherapy or in combination with oral glucose-lowering agents), mean ± standard deviation (SD) or median (interquartile range or IQR) age, duration of diabetes, SBP, DBP, BMI, FBG, HbA1c, prevalence and mode of diagnosis of the diabetes complication of interest, and the related predictors. The five chronic microvascular and macrovascular diabetes complications of interest were diabetic neuropathy, nephropathy, retinopathy, diabetic foot disease (DFD), and peripheral arterial disease (PAD).

### Operational definitions

For the chronic diabetes complications of interest, we considered diagnoses made based on self-report, documentation in medical records, or standard recommended diagnostic approaches. For the latter approach of diagnosing the diabetes complications, we considered the presence of albuminuria (>30 mg/g or 3 mg/mmol) and/or an estimated glomerular filtration rate (e-GFR) of <60 ml/min/1.73 m2 for the diagnosis of diabetic nephropathy [10]. For the diagnosis of diabetic peripheral neuropathy, we considered the presence of suggestive symptoms such as intense pain, paraesthesia, and numbness in the feet, features on neurological examination such as loss of pain and fine touch sensation, reduced ankle reflexes, and the use of neuropathy grading scores such as neuropathy symptom score (NSS) and neuropathy disability score (NDS) [11]. Diabetic retinopathy was diagnosed based on a fundoscopic examination demonstrating the presence of suggestive features such as microaneurysms, retinal dot and blot haemorrhages, hard exudates or cotton wool spots, neovascularisation, vitreous or pre-retinal haemorrhage, retinal detachment, and diabetic macular oedema [12]. The presence of peripheral neuropathy, PAD, infection, ulcer(s), neuro-osteoarthropathy, gangrene, or amputation was suggestive of DFD [13]. A diagnosis of PAD was made based on an ankle-brachial index of <0.90 on arterial Doppler ultrasonography and/or the presence of clinical features such as hyperpigmentation of the skin of the feet, loss of hair, and feeble or absent foot pulses [14].

### Assessment of the quality and publication bias of studies

The quality of all eligible studies included in the systematic review and meta-analysis was assessed using the modified Newcastle-Ottawa Scale (NOS) [15]. This was done independently by one author (APK).

### Study outcomes

The study outcomes were the prevalence and predictors of five chronic microvascular and macrovascular diabetes complications.

### Data analysis

All analyses were performed using STATA MP 18.0 statistical software (Stata Corp, College Station, Texas USA). The results were reported in line with PRISMA 2000 guidelines. The descriptive data of all eligible studies were summarised based on the type of variable reported. For categorical variables, a meta-analysis of proportions was performed using the Freeman-Tukey double arcsine transformation to stabilize variances. We calculated a weighted mean of means for continuous variables reported as means, with weights proportional to study sample sizes. When medians were reported, we pooled the medians by calculating the median of medians. The pooled prevalence of each diabetes complication was determined using a random effect model meta-analysis of proportions using the Freeman-Tukey double arcsine transformation and presented in the form of forest plots.

The heterogeneity of the included studies was assessed using I^2^ values. Based on the Cochrane collaboration guide, the I^2^ values of 0%–40%, 30%–60%, 50%–90%, and 75%–100% were considered not important, moderate, substantial, and considerable levels of heterogeneity, respectively [16]. We conducted univariate random-effects meta-regression to explore potential sources of heterogeneity and identify study-level covariates that may influence the variability in prevalence estimates across studies. We assessed the presence of publication bias visually using funnel plots and statistically using the Egger test of bias with p<0.05 indicating significant publication bias [17]. The information about the included studies was summarised in tables.

### Ethical approval

Because this was a systematic review and meta-analysis of published studies, no prior ethical approval was required.

## RESULTS

Figure 1 summarises the article selection in a PRISMA flow diagram.

**Figure 1.**
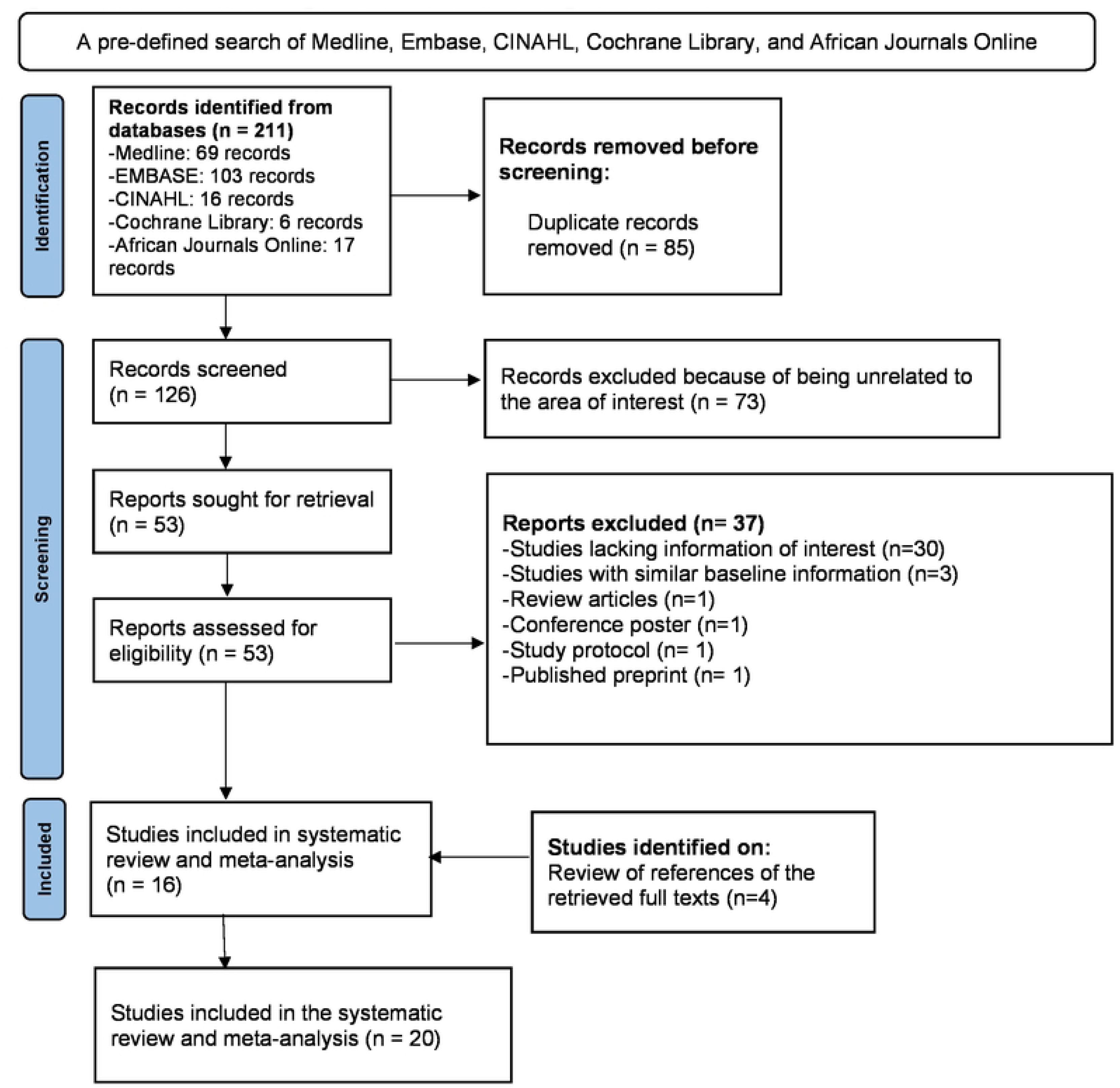
PRISMA flow diagram of selection of eligible studies.

The literature search returned a total of 211 articles. From these, 85 duplicates were removed. We reviewed the titles and abstracts of the remaining 126 articles and 53 articles were identified for full-text retrieval. Of these, 37 were excluded, with 16 articles remaining. On hand-searching the references of the retrieved full texts, four additional articles were identified to make a total of 20 articles included in this systematic review and meta-analysis.

### Overall characteristics of participants included in this systematic review and meta-analysis

Table 1 summarises the overall sociodemographic, clinical, anthropometric, and metabolic characteristics of all study participants.

**Table 1.** Overall characteristics of the participants included in the systematic review and meta-analysis.

| Characteristics |  | Pooled value | Number of studies |
| --- | --- | --- | --- |
| **Demographic and clinical** |  |  |  |
| Age, years | Mean (SD) | 54.8 (3.6) | 16 |
| Female participants | %, 95% CI | 61.1 (57.1 – 65.2) | 20 |
| Current smoking history | %, 95% CI | 9.3 (4.3 – 14.4) | 12 |
| Family history of diabetes | %, 95% CI | 41.3 (15.8 – 66.7) | 4 |
| Co-existing HIV | %, 95% CI | 9.9 (5.5 – 14.2) | 6 |
| Co-existing hypertension | %, 95% CI | 51.9 (42.8 – 60.9) | 13 |
| Insulin therapy (monotherapy or in combination with oral glucose-lowering agents) | %, 95% CI | 38.4 (27.2 – 49.6) | 9 |
| Duration of diabetes, years | Mean (SD) | 6.9 (1.6) | 7 |
|  | Median (IQR) | NA | 2 |
| Systolic blood pressure, mmHg | Mean (SD) | 139.5 (1.5) | 3 |
|  | Median (IQR) | 140 (125 – 145) | 3 |
| Diastolic blood pressure, mmHg | Mean (SD) | 84.9 (4.0) | 3 |
|  | Median (IQR) | 81 (80 – 84) | 3 |
| **Anthropometric** |  |  |  |
| Body mass index, kg/m<sup>2</sup> | Mean (SD) | 26.2 (1.4) | 6 |
|  | Median (IQR) | NA | 2 |
| **Metabolic** |  |  |  |
| Fasting blood glucose, mmol/l | Mean (SD) | 10.5 (0.8) | 3 |
|  | Median (IQR) | NA | 2 |
| Glycated haemoglobin, % | Mean (SD) | 10.5 (2.2) | 4 |
|  | Median (IQR) | 9.1 (8.1 – 10.6) | 3 |
| Glycated haemoglobin, mmol/mol | Mean (SD) | 91.0 (1.0) | 4 |
|  | Median (IQR) | 76.0 (65.0-92.0) | 3 |

CI- Confidence interval, IQR- Interquartile range, NA- Not applicable, SD- Standard deviation

A total of 20 studies involving 11,400 participants (ranging from 41 to 5,730 participants) were included in this systematic review and meta-analysis. The pooled mean (SD) age, duration of diabetes, and HbA1c of the participants were 54.8 (3.6) years, 6.9 (1.6) years, and 10.5 (2.2)% or 91.0 (1.0) mmol/mol respectively. About 61.1% (95% confidence interval [CI] 57.1-65.2) of the participants were female. The pooled prevalence of co-existing hypertension and HIV was 51.9% (95% CI 42.8-60.9) and 9.9% (95% CI 5.5-14.2), respectively.

### Characteristics of studies included in this systematic review and meta-analysis

Table 2 summarises the characteristics of the included studies.

**Table 2.** Characteristics of each study included in the systematic review and meta-analysis.

| **Last name of 1<sup>st</sup> author, year of publication, and number of participants** | **Sociodemographic characteristics** | **Clinical and anthropometric characteristics** | **Diabetes complication(s) screened and mode of diagnosis** | **Prevalence and predictors of the screened diabetes complication(s)** |
| --- | --- | --- | --- | --- |
| Alimwenda et al 2022<br>n = 1000 participants | -Mean age: 51.1 ± 13.1 years<br>-Female participants: 674 (67.4%)<br>-History of current smoking: 20.8% | -Co-existing HIV: 16.9% | -Diabetic neuropathy, nephropathy, foot disease, and retinopathy (all self-reported) | -Diabetic neuropathy: 54.0%<br>-Diabetic nephropathy: 15.6%<br>-Diabetic foot disease: 11.0%<br>-Diabetic retinopathy: 9.0% |
| Arunga et al 2020<br>n = 5,730 participants | -Median (IQR) age: 56.0 (46.0-100.0) years<br>-Female participants: 4,189 (73.1%) |  | -Diabetic retinopathy (diagnosed based on findings of retinal photography using a portable non-mydratic fundus camera) | -Diabetic retinopathy: 5.1% |
| Bateganya et al 2003<br>n = 120 participants | -Mean age: 49.0 ± 18.0 years<br>-Female participants: 69 (57.5%)<br>-History of current smoking: 11.7% | -Co-existing HT: 36.7%<br>-Participants on insulin therapy: 47.5% | -Diabetic neuropathy (diagnosed based on the presence of suggestive symptoms and clinical examination)<br>-Diabetic nephropathy (diagnosed based on the presence of proteinuria)<br>-Diabetic foot disease (diagnosed based on clinical examination) | -Diabetic neuropathy: 38.5%<br>-Diabetic nephropathy: 25.8%<br>-Diabetic foot disease: 3.3% |
| Kibirige et al 2014<br>n = 250 participants | -Mean age: 51.6 ± 9.2 years<br>-Female participants: 155 (62.0%)<br>-Family history of diabetes: 37.2%<br>-History of current smoking: 1.2% | -Mean BMI: 24.5 ± 3.9 kg/m<sup>2</sup><br>-Co-existing HT: 78.4%<br>-Participants on metformin and sulfonylureas: 71.2%<br>-Participants on metformin only: 28.8%<br>-Participants on insulin therapy: 20% | -Diabetic neuropathy, nephropathy, retinopathy, foot disease, PAD, and ischaemic heart disease (all self-reported) | -Diabetic neuropathy: 31.2%<br>-Diabetic retinopathy: 9.6%<br>-Diabetic foot disease: 3.2%<br>-PAD: 2.0%<br>-Ischaemic heart disease: 1.2%<br>-Diabetic nephropathy: 0.8% |
| Kibirige et al 2023<br>n = 519 participants | -Median (IQR) age: 48 (39-57) years<br>-Female participants: 292 (56.3%) | -Co-existing HT: 33.7%<br>-Median (IQR) SBP: 125 (115-136) mmHg<br>-Median (IQR) DBP: 84 (77-91) mmHg | -Diabetic nephropathy (diagnosed based on the presence of albuminuria) | -Diabetic nephropathy: 33.7%.<br>**<u>Predictors</u>** |
| | | -Median (IQR) BMI: 27.4 (23.4-31.4) mmHg<br>-Median (IQR) FBG: 8.6 (6.3-13.4) mmol/l<br>-Median (IQR) HbA1c: 10.6 (7.8-12.5) % or 92.0 (62.0-113.0) mmol/mol | | -Self-reported HT comorbidity (AOR: 1.76, 95% CI 1.24-2.48, p=0.002)<br>-BMI ≥30 kg/m<sup>2</sup> (AOR: 0.61, 95% CI 0.41-0.91, p= 0.02) |
| Kiconco et al 2019<br>n = 140 participants | -Female participants: 95 (67.9%)<br>-Family history of diabetes: 77.9%<br>-History of current smoking: 2.1% | -Mean duration of diabetes: 6.8 years<br>-Co-existing HT: 42.1%<br>-Mean SBP: 138.6 mmHg<br>-Mean DBP: 78.1 mmHg<br>-Mean BMI: 24.4 kg/m<sup>2</sup><br>-Mean ± SD FBG: 9.3 ± 5.4 mmol/l | -Diabetic nephropathy (diagnosed based on the presence of microalbuminuria) | -Diabetic nephropathy: 23.6%<br>**Predictors**<br>Family history of diabetes ($\beta$= 0.275, 95% CI 0.043-0.508, p=0.002) |
| Kisozi et al 2017<br>n = 248 participants | -Mean age: 48.5 ± 13.4 years<br>-Female participants: 94 (37.9%) | | -Diabetic neuropathy (diagnosed based on the presence of suggestive symptoms and the NSS and NDS) | -Diabetic neuropathy: 29.4%<br>**Predictors**<br>-Age >60 years (AOR: 3.72, 95%CI 1.25-11.03, p=0.018)<br>-History of a foot ulcer (AOR- 2.59, 95% CI 1.03-6.49, p=0.042) |
| Lumori et al 2022<br>n = 195 participants | -Mean age: 62.0 ± 11.5 years<br>-Female participants: 141 (72.3%)<br>-Current history of smoking: 0.5% | -Median (IQR) duration of diabetes: 10.0 (7.0-15.0) years<br>-Co-existing HT: 74.9%<br>-Co-existing HIV: 7.2%<br>-Median (IQR) SBP: 145 (128-158) mmHg<br>- Median (IQR) DBP: 81 (75-89) mmHg<br>-Median (IQR) HbA1c: 9.1 (7.7-10.9) % or 76.0 (61.0-96.0) mmol/mol | -Diabetic neuropathy (diagnosed based on the presence of suggestive symptoms) | -Diabetic neuropathy: 61.0% |
| Magan et al 2019<br>n = 41 participants | -Mean ± SD age: 50.4 ± 12.5 years<br>-Female participants: 26 (63.4%) | -Mean ± SD duration of diabetes: 8.1 ± 6.4 years<br>-Co-existing HT: 65.9%<br>-Participants on insulin therapy: 51.2% | -Diabetic retinopathy (diagnosed based on findings of indirect fundoscopy ± retinal photography using a portable non-mydratic fundus camera) | -Diabetic retinopathy: 19.5% |
| Migisha et al 2020<br>n = 299 participants | -Mean ± SD age: 50.1 ± 9.8 years<br>-Female participants: 208 (69.6%)<br>-Current history of smoking: 25.7% | -Mean ± SD duration of diabetes: 5.8 ± 5.9 years<br>-Mean ± SD BMI: 27.4 ± 5.6 kg/m<sup>2</sup><br>-Mean ± SD SBP: 141.7 ± 22.6 mmHg | -Diabetic neuropathy and retinopathy (all self-reported) | -Diabetic neuropathy: 61.2%<br>-Diabetic retinopathy: 22.7% |
| | | -Mean ± SD DBP: 87.1 ± 10.7 mmHg<br>-Mean ± SD FBG: 11.2 ± 4.8 mmHg<br>-Mean ± SD HbA1c: 9.7 ± 2.6% or 83.0 ± 5.0 mmol/mol | | |
| Muddu et al 2018<br>n = 175 participants | -Mean ± SD age: 46.0 ± 15.0 years<br>-Female participants: 85 (48.6%) | -Mean ± SD HbA1c: 13.9 ± 5.3% or 128.0 ± 34.0 mmol/mol | -Diabetic nephropathy (diagnosed based on the presence of microalbuminuria) | -Diabetic nephropathy: 47.4%<br>**Predictors**<br>-Pregnancy (AOR: 7.74, 95% CI 1.01-76.47, p=0.05)<br>-Mild and moderate physical activity (AOR: 0.08, 95% CI 0.01-0.95, p=0.046) |
| Munyambalu et al 2023<br>n = 319 participants | -Mean age: 59.4 ± 14.6 years<br>-Female participants: 197 (61.8%)<br>-History of current smoking: 13.5% | -Mean ± SD duration of diabetes: 7.3 ± 6.4 years<br>-Co-existing HT: 50.2%<br>-Co-existing HIV: 9.1%<br>-Participants on insulin therapy: 12.9%<br>-Mean ± SD BMI: 26.3 ± 3.5 years<br>-Mean ± SD SBP: 138.4 ± 19.8 mmHg<br>-Mean ± SD DBP: 85.8 ± 13.0 mmHg<br>-Mean ± SD FBG: 10.4 ± 5.2 mmol/l | -Diabetic neuropathy (diagnosed based on the presence of suggestive symptoms and the NDS) | -Diabetic neuropathy: 65.8% |
| Mwebaze et al 2014<br>n = 146 participants | -Mean ± SD age: 53.9 ± 12.4 years<br>-Female participants: 71 (48.6%)<br>-History of current smoking: 0.7% | -Mean ± SD duration of diabetes: 5.8 ± 3.2 years<br>-Co-existing HT: 47.3%<br>-Participants on insulin therapy: 36.7%<br>-Mean ± SD HbA1c: 9.5 ± 2.5% or 80.0 ± 4.0 mmol/mol | -PAD (diagnosed based on an ABI ≤0.90 on Doppler ultrasonography) | -PAD: 39% |
| Nambuya et al 1996<br>n = 252 participants | -Mean age: 45 years<br>-Female participants: 135 (53.6%)<br>-Family history of diabetes: 16.3% | -Co-existing HT: 27.3%<br>-Participants on insulin therapy: 52.8% | -Diabetic neuropathy, nephropathy, foot disease, and ischaemic heart disease (all self-reported) | -Diabetic neuropathy: 46.4%<br>-Diabetic nephropathy: 4.8%<br>-Ischaemic heart disease: 4.8%<br>-Diabetic foot disease: 4.0% |
| Okello et al 2014<br>n = 229 participants | -Median (IQR) age: 60 (55-66) years<br>-Female participants: 146 (63.7%) | -Median (IQR) duration of diabetes: 1.0 (0.3-2.3) years<br>-Co-existing HT: 49.3%<br>-Co-existing HIV: 7.9% | -PAD (diagnosed based on an ABI ≤0.90 on doppler ultrasonography) | -PAD: 24%<br>**Predictors**<br>-Female sex (AOR: 2.25, 95% CI 1.06-4.77, p=0.034) |
|  | -Family history of diabetes: 34.1%<br>-History of current smoking: 5.2% | -Participants on metformin only: 51.6%<br>-Participants on metformin and glibenclamide: 15.7%<br>-Participants on insulin therapy: 29.3%<br>-Median (IQR) SBP: 140 (124-160) mmHg<br>-Median (IQR) DBP: 80 (70-90) mmHg<br>-Median (IQR) FBG: 8.6 (6.6-13.7) mmol/l<br>-Median (IQR) HbA1c: 8.1 (6.7-10.1) % or 65.0 (50.0-87.0) mmol/mol |  | -Current high blood pressure (AOR: 2.59, 95% CI 1.25-5.33, p=0.01)<br>-Using glibenclamide (AOR: 3.47, 95% CI 1.55-7.76, p=0.002) |
| Pario et al 2022<br>n = 60 participants | -Female participants: 42 (70%) | -Participants on insulin therapy: 33.3% | -Diabetic neuropathy (diagnosed based on the presence of suggestive symptoms)<br>-PAD (diagnosed based on the presence of symptoms of claudication and rest pain) | -Diabetic neuropathy: 91.7%<br>-PAD: 65% |
| Patrick et al 2021<br>n = 223 participants | -Mean age: 61.1 ± 4.0 years<br>-Female participants: 153 (68.9%)<br>-History of current smoking: 2.2% | -Mean duration of diabetes: 6.1 ± 7.1 years<br>-BMI: 27.9 ± 5.2 kg/m<sup>2</sup> | -Diabetic neuropathy and retinopathy (all self-reported) | -Diabetic neuropathy: 85.7%<br>-Diabetic retinopathy: 65.1% |
| Sikhondze et al 2022<br>n = 62 participants | -Mean ± SD age: 57.0 ± 12.3 years<br>-Female participants: 35 (56.5%)<br>-History of current smoking: 22.6% | -Co-existing HT: 40.3%<br>-Co-existing HIV: 17.7% | -PAD (diagnosed based on an ABI ≤0.90 on Doppler ultrasonography)<br>-Diabetic nephropathy and retinopathy (self-reported) | -PAD: 35.5%<br>-Diabetic retinopathy: 6.5%<br>-Diabetic nephropathy: 3.2% |
| Tino et al 2020<br>n = 1,275 participants | -Median (IQR) age: 54 (44-65) years<br>-Female participants: 770 (60.4%)<br>-History of current smoking: 10.9% | -Co-existing HT: 69.6% | -Diabetic neuropathy, nephropathy, and PAD (all self-reported) | -Diabetic neuropathy: 37.7%<br>-Diabetic nephropathy: 6.4%<br>-PAD: 2.3% |
| Vahwere et al 2023<br>n = 117 participants | -Mean age: 57.1 years<br>-Female participants: 72 (61.5%) | -Mean ± SD duration of diabetes: 10.9 ± 9.7 years<br>-Co-existing HT: 59%<br>-Participants on insulin therapy: 65.0%<br>-Mean ± SD BMI: 25.3 ± 5.9 mmol/l | -Diabetic neuropathy (diagnosed based on the presence of suggestive symptoms)<br>-PAD (diagnosed based on the presence of symptoms of claudication, features of gangrene, and an ABI | -Diabetic neuropathy: 79.5%<br>-PAD: 30.7% |
| | | -Mean ± SD HbA1c: 8.6 ± 2.6% or 70.0 ± 5.0 mmol/mol | ≤0.90 on Doppler ultrasonography) | |

ABI- Ankle brachial index, AOR- Adjusted odds ratio, BMI- Body mass index, CI- Confidence interval, DBP- Diastolic blood pressure, FBG- Fasting blood glucose, HbA1c- Glycated haemoglobin, HT- Hypertension, IQR- Interquartile range, NDS- Neuropathy disability score, NSS- Neuropathy symptom score, PAD- Peripheral arterial disease, SBP- Systolic blood pressure, SD- Standard deviation

A total of 20 studies were included in this systematic review and meta-analysis [18–37]. The majority of the studies were cross-sectional in design (17 studies, 85%) [19, 20, 22–35, 37] with only three studies being retrospective [18, 21, 36]. Of these studies, 15 (75%) were conducted in public hospitals [18–20, 23–35] while three (15%) were conducted in private hospitals [21, 29, 36]. Two studies were conducted in both public and private hospitals [22, 37]. Considerable heterogeneity was noted in the analysis of studies with the I^2^ ranging from 90.22% to 99.60%.

### Assessment of study quality

The assessment of the quality of studies is summarised in Supplementary Table 2. Based on the modified NOS, all studies were of low quality. Of the 20 studies, 14 (70%) were considered satisfactory while the remaining six studies were considered unsatisfactory.

### Assessment of the publication bias of the studies

The funnel plots assessing publication bias are summarised in Supplementary Figures 1-5. The funnel plot symmetry showed no evidence of publication bias in the studies assessing the prevalence of diabetic neuropathy (**Egger p-value = 0.655, Supplementary Figure 1**), diabetic retinopathy (**Egger p-value = 0.434, Supplementary Figure 2**), and diabetic foot (**Egger p-value = 0.280, Supplementary Figure 5**). However, the funnel plot was asymmetrical for the meta-analysis of studies assessing the prevalence of diabetic nephropathy (**Egger p-value = 0.005, Supplementary Figure 3**) and PAD (**Egger p-value<0.001, Supplementary Figure 4**), showing evidence of publication bias. Non-parametric trim and fill analysis for publication bias for both complications did not result in any differences in the prevalence, whether with right or left imputation.

### Prevalence of chronic diabetes complications

The pooled prevalence of the five chronic microvascular and macrovascular diabetes complications is summarised as forest plots in Figures 2-6.

**Figure 2.**
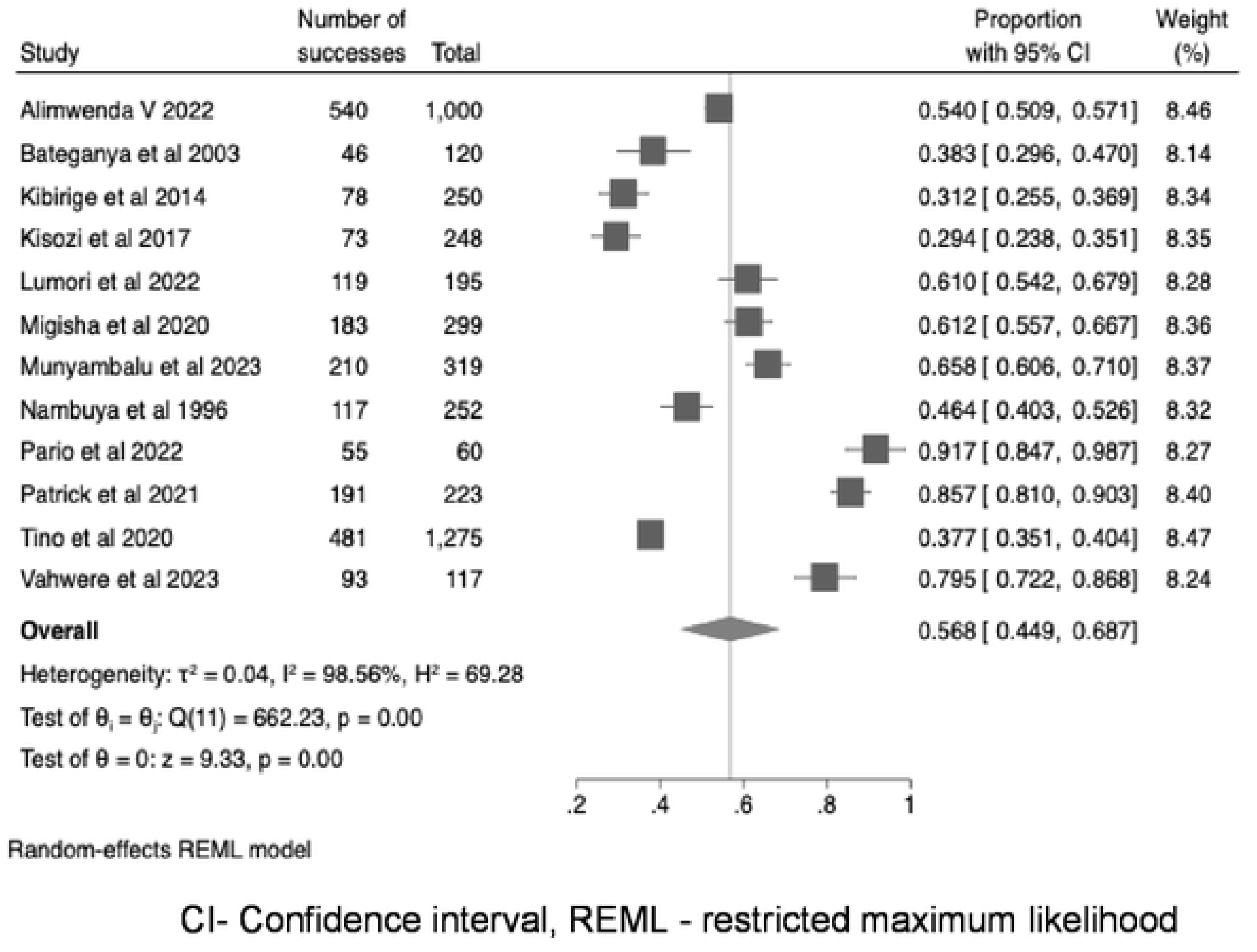
The pooled prevalence of diabetic neuropathy.

Regarding chronic microvascular diabetes complications, information on the prevalence of diabetic neuropathy, nephropathy, and retinopathy was reported by 12 studies [18, 20, 21, 24, 25, 28, 29, 31, 33, 34, 36, 37], nine studies [18, 20–23, 27, 31, 35, 36], and seven studies [18, 19, 21, 26, 34–36], respectively. For chronic macrovascular diabetes complications, information on the prevalence of PAD and DFD was reported by six studies [21, 30, 32, 33, 35, 37] and four studies [18, 20, 21, 31], respectively.

Of the 12 studies, the prevalence of diabetic neuropathy was based on self-report (presence of suggestive symptoms only) in seven studies (58.3%) [18, 21, 25, 31, 34, 36, 37]. The remaining studies diagnosed diabetic neuropathy based on the presence of suggestive symptoms, a neurological clinical examination, and the use of neuropathy grading scores such as NDS and NSS [20, 24, 28, 29, 33]. The pooled prevalence of diabetic neuropathy in the 12 studies was 56.8% (95% CI 44.9-68.7, I^2^ = 98.56%, p<0.001) (Figure 2).

Of the seven studies reporting information on the prevalence of diabetic retinopathy, only two studies (28.6%) made the diagnosis based on fundoscopic examination [19, 26]. The rest of the studies based the diagnosis on self-report [18, 21, 34–36]. The pooled prevalence of diabetic retinopathy in the seven studies was 19.5% (95% CI 3.9-35.2, I^2^ = 99.60%, p<0.001) (Figure 3).

**Figure 3.**
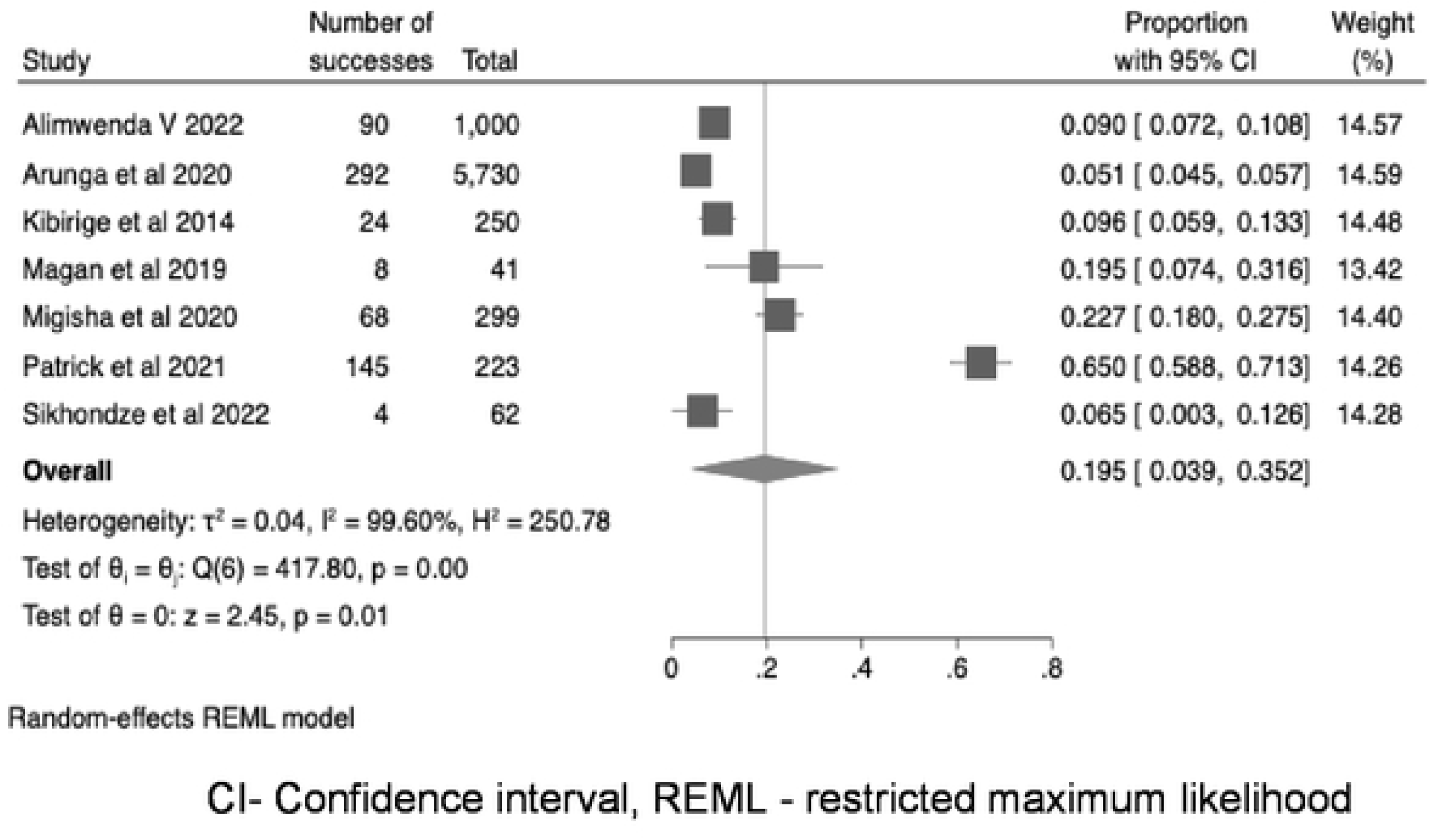
The pooled prevalence of diabetic retinopathy.

Among the nine studies that reported the prevalence of diabetic nephropathy, five (55.6%) based the diagnosis on the presence of albuminuria [20, 22, 23, 27, 31]. The diagnosis in the remaining studies was based on self-report [18, 21, 35, 36]. The pooled prevalence of diabetic nephropathy in these studies was 17.7 (95% CI 7.3- 28.0, I^2^ = 99.36%, p<0.001) (Figure 4).

**Figure 4.**
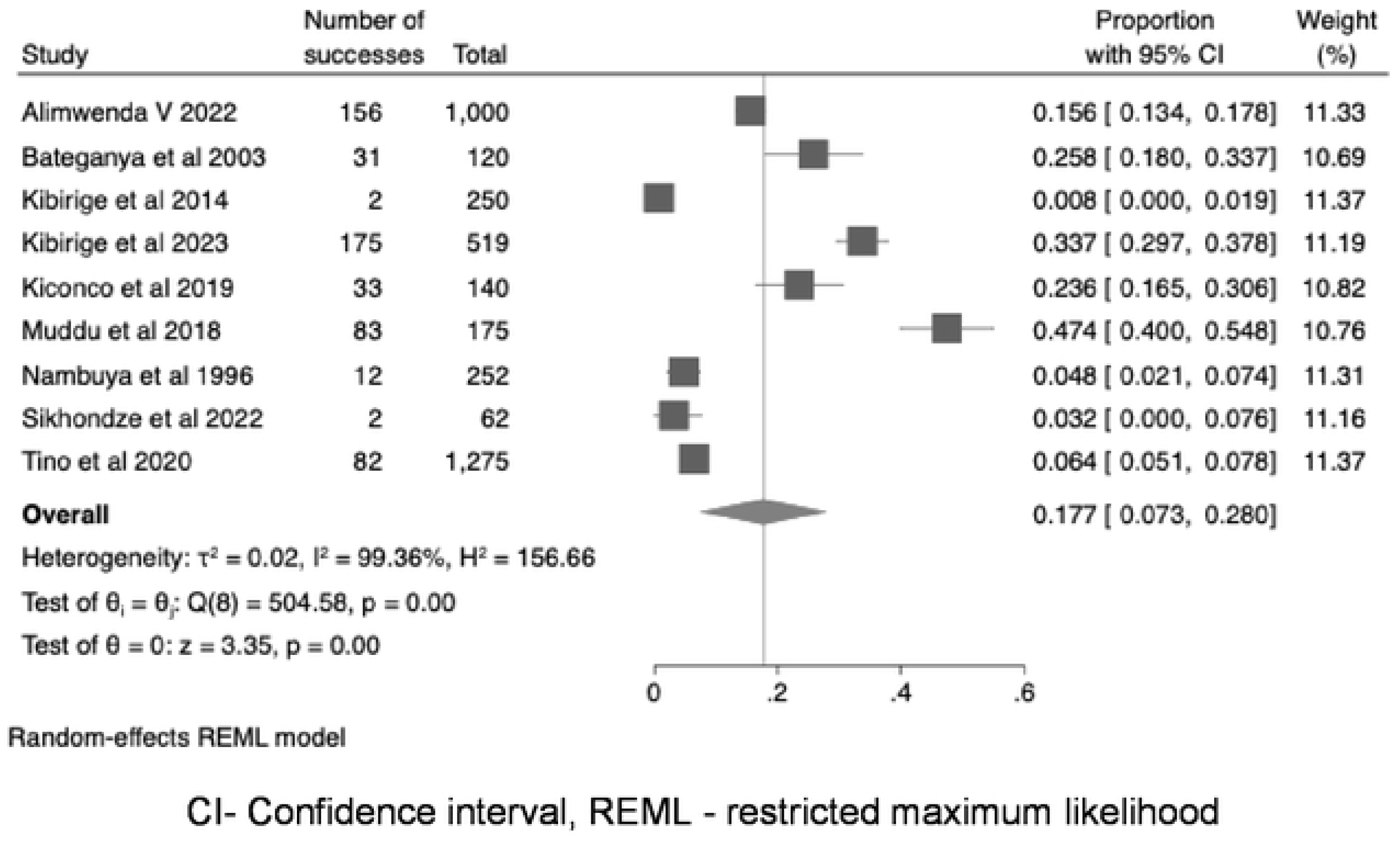
The pooled prevalence of diabetic nephropathy.

For the macrovascular diabetes complications, regarding the prevalence of PAD reported by the six studies, the diagnosis was made based on clinical examination and/or Doppler ultrasonography in five studies (83.3%) [30, 32, 33, 35, 37], with one study basing the diagnosis on self-report [21]. The pooled prevalence of PAD was 32.2 (95% CI 15.8-48.7, I^2^ = 97.67%, p<0.001) (Figure 5).

**Figure 5.**
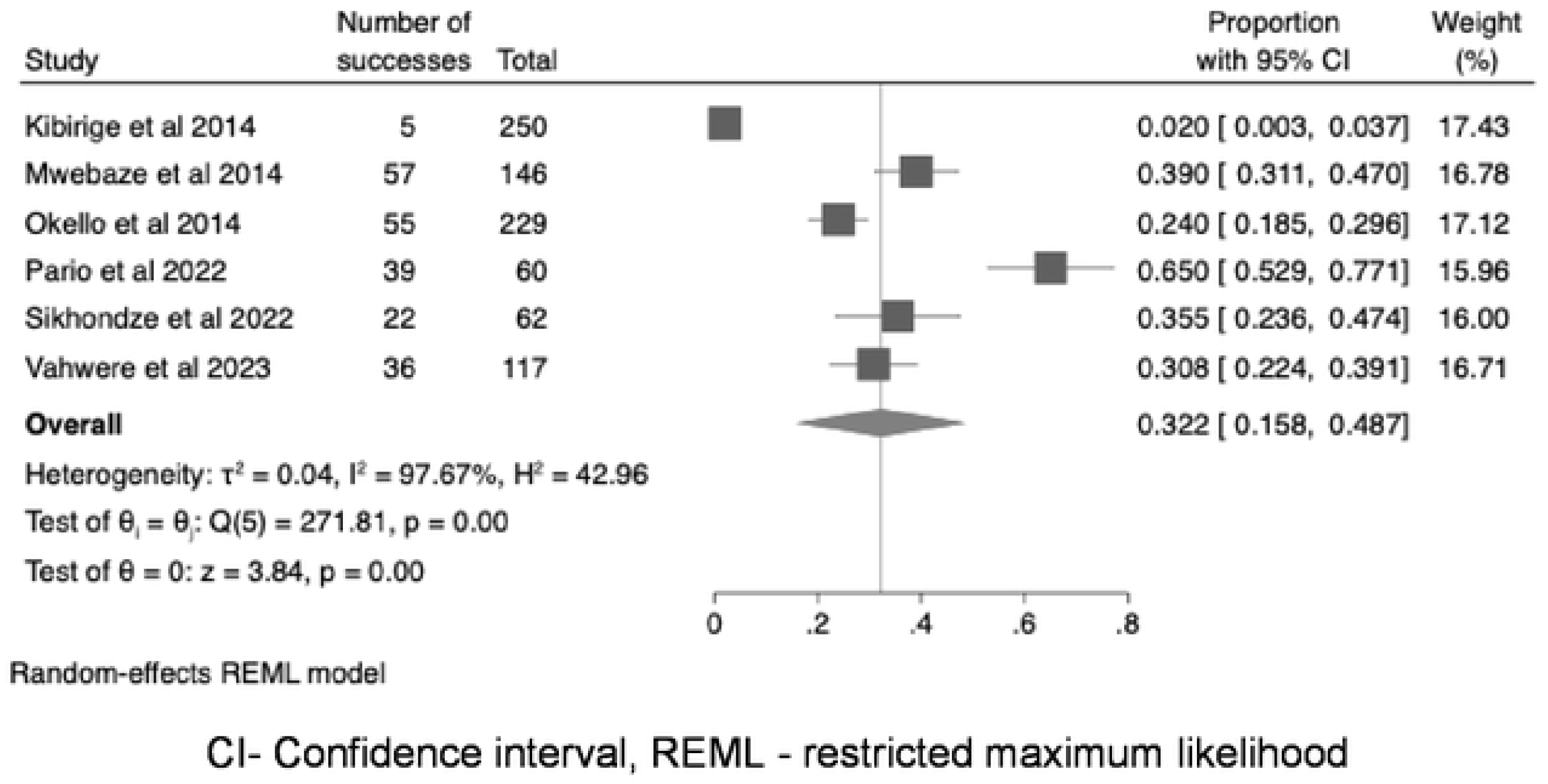
The pooled prevalence of peripheral arterial disease.

The diagnosis of DFD was based on clinical examination in all four studies [18, 20, 21, 31], documenting a pooled prevalence of 5.5% (95% CI 1.7-9.2, I^2^= 90.22%, p<0.001) (Figure 6).

**Figure 6.**
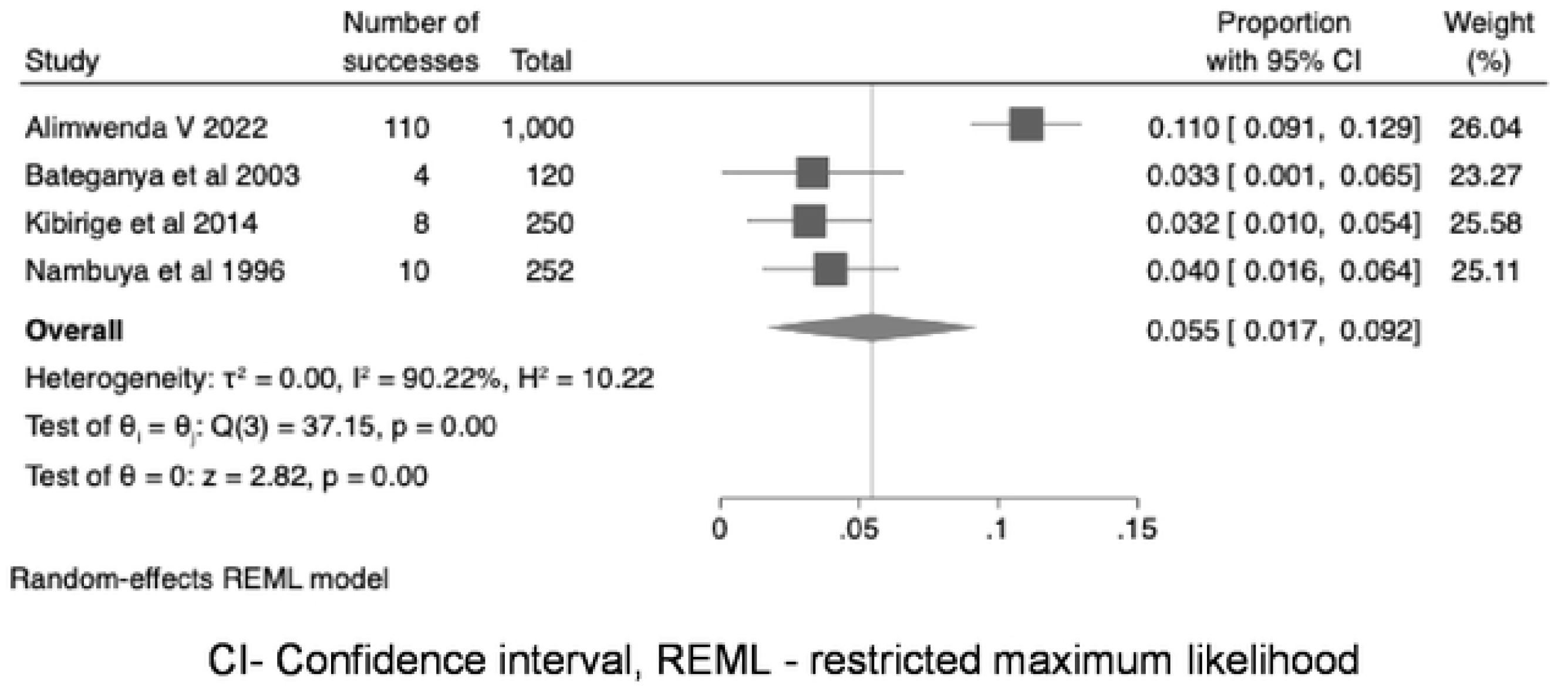
The pooled prevalence of diabetic foot disease.

### Significant predictors of chronic diabetes complications

The information on the significant predictors of chronic diabetes complications was reported by only five studies (25%) [22–24, 27, 32]. Of these, three studies reported predictors of diabetic nephropathy [22, 23, 27] while the remaining two studies reported predictors of diabetic neuropathy [24] and PAD [32].

The predictors of diabetic nephropathy in the three studies were self-reported hypertension comorbidity (adjusted odds ratio [AOR]: 1.76, 95% CI 1.24-2.48, p=0.002), pregnancy (AOR: 7.74, 95% CI 1.01-76.47, p=0.05), and family history of diabetes (β= 0.275, 95% CI 0.043-0.508, p=0.002) which were associated with increased odds of having diabetic nephropathy, while BMI ≥30 kg/m^2^ (AOR: 0.61, 95% CI 0.41-0.91, p=0.02) and mild to moderate physical activity (AOR: 0.08, 95% CI 0.01- 0.95, p=0.046) reduced the odds [22, 23, 27].

In the study by Kisozi et al, the predictors of diabetic neuropathy were a history of a foot ulcer (AOR: 2.59, 95% CI 1.03-6.49, p=0.042) and age >60 years (AOR: 3.72, 95% CI 1.25-11.03, p=0.018) [24]. The predictors of PAD in the study by Okello et al were female sex (AOR: 2.25, 95% CI 1.06-4.77, p=0.034), current hypertension (AOR: 2.59, 95% CI 1.25-5.33, p=0.01), and being on glibenclamide (AOR: 3.47, 95% CI 1.55-7.76, p=0.002) [32].

### Meta-regression analysis

The meta-regression models explored the influence of various covariates such as age, proportion of females, smoking status, co-existing HIV, insulin therapy, hypertension, and BMI on the proportion of individuals affected by these complications (**Supplementary Table 3)**.

For diabetic neuropathy, the mean age and proportion of females were found to be a significant predictor of neuropathy prevalence. Each unit increase in mean age in years was associated with a 2.34% increase in the prevalence of neuropathy (coefficient: 0.0234; 95% CI: 0.0079, 0.0388; p = 0.003). The model explained 47.6% heterogeneity (R^2^ = 47.6%). Similarly, each percentage increase in the proportion of females in the study was significantly associated with a 1.41% increase in the prevalence of neuropathy (coefficient: 0.0141; 95% CI: 0.0034, 0.0249; p = 0.010, R^2^ = 34.8%). No other variables, including smoking status, coexisting HIV, insulin usage, and hypertension, were significant predictors (p > 0.05).

For PAD, the proportion of patients with hypertension was the only significant predictor, with each unit increase in hypertension prevalence associated with a 0.92% decrease in PAD prevalence (coefficient: −0.0092; 95% CI: −0.0142, −0.0041; p = 0.000), explaining 77.64% of the variance. Other variables, including mean age, proportion of females, smoking status, and insulin usage, were not statistically significant (p >0.05).

For all other diabetes complications, no significant factors were found to be associated with their prevalence.

## DISCUSSION

In this systematic review and meta-analysis, we report a high prevalence of chronic diabetes complications, especially diabetic neuropathy and PAD, in adult Ugandans with diabetes. Generally, several studies, systematic reviews, and meta-analyses have reported a high prevalence of diabetes complications in adult African populations with diabetic neuropathy and retinopathy being the most prevalent [3, 38–43].

In one systematic review and meta-analysis of 23 studies describing 269,691 Africans with diabetes by Shiferaw et al, a comparably high prevalence of diabetic neuropathy of 46% was reported [43]. Regarding the prevalence of diabetic retinopathy, several studies conducted in Nigeria [44], Botswana [45], Ethiopia [46, 47], and South Africa [48] have reported findings comparable to our study with the prevalence ranging between 17.7% to 21%. The prevalence of diabetic nephropathy in adult Africans reported by some systematic reviews and meta-analyses varies widely in comparison to our study findings. This heterogeneity in the prevalence may be explained by differences in study definitions of diabetic nephropathy (self-reported vs. diagnosed based on assessment for albuminuria and/or e-GFR). The systematic review and meta-analysis by Kibirige et al [3] and Wagnew et al [49] reported a higher prevalence of diabetic nephropathy in large adult African populations with diabetes of 31% and 35.3%, respectively, compared with what we observed in our study (18%).

Regarding macrovascular diabetes complications, a high prevalence of PAD, similar to what we observed in our study, has been reported in adult Africans with diabetes [50–53]. Studies conducted in Nigeria [51], Ethiopia [52], and Egypt [53] reported a high comparable prevalence of PAD ranging between 30.7% to 38.5%.

As observed in our study, DFD has been reported as the least prevalent diabetes complication in adult Africans with diabetes in systematic reviews and meta-analyses. Kibirige et al [3] and Rigato et al [42] reported a prevalence of DFD in large adult African populations of 11% and 13%, respectively.

There are several plausible explanations for the high prevalence of these chronic diabetes complications in adult Ugandans with diabetes. The rate of undiagnosed diabetes remains very high, with the majority of people presenting late with diabetes complications [4, 5]. Glycaemic, blood pressure, lipid screening, management, and monitoring in most adult Ugandans with diabetes remains largely suboptimal [21, 54, 55]. This significantly increases the risk of onset and progression of chronic diabetes complications.

Additionally, Uganda still grapples with the challenges of low access to affordable essential diabetes medicines and diagnostic tests, gaps in knowledge in preventing and managing diabetes among healthcare professionals and individuals living with diabetes, and a poorly structured healthcare system to manage chronic conditions such as diabetes [7, 8, 55–58].

Regarding the predictors of diabetes complications, consistent with findings from other studies, co-existing hypertension [41, 49] and reduced physical activity [59] have been associated with increased odds of developing diabetic nephropathy. While obesity has been widely reported to increase the risk of diabetic nephropathy in most populations [60], it was associated with a reduced risk in one of the studies that we included in the meta-analysis [22]. A similar protective effect of obesity towards developing diabetic nephropathy was also reported in a study conducted in rural South Africa. In this study, severe obesity (defined as BMI >33 kg/m^2^) reduced the risk of developing microalbuminuria by 73% [61]. This demonstrates differences in some of the risk factors for diabetic nephropathy across populations.

Similar to the Ugandan study by Kisozi et al [24], one study conducted on adult Ethiopians with type 2 diabetes reported an association between age >60 years and the presence of diabetic neuropathy [62]. Additionally, as reported in another Ugandan study by Okello et al [32], hypertension was identified as one of the predictors of PAD in a Southern African [63] and Central African population [64].

### Strengths and limitations

To our knowledge, this is the first systematic review and meta-analysis to document the prevalence of microvascular and macrovascular diabetes complications in adult Ugandans with diabetes. The studies were conducted in the different regions of Uganda, improving the generalisability of the results of the systematic review and meta-analysis.

Despite these strengths, the systematic review and meta-analysis had some limitations. Some of the included studies used self-reporting as the basis for diagnosing the chronic diabetes complication of interest. This is subject to recall bias and reporting imprecise results. There was considerable heterogeneity in the study findings possibly due to the differences in screening procedures, study definitions for the complications of interest, and study settings (public vs. private healthcare facilities). The studies included in this systematic review and meta-analysis were of low-quality rating.

### Conclusions

This systematic review and meta-analysis reported a high prevalence of microvascular and macrovascular diabetes complications in an adult Ugandan population with diabetes. There is a need to improve the early identification of individuals with undiagnosed diabetes in routine care in Uganda. Early routine screening and optimal management of diabetes complications in adult individuals with diabetes should be strongly encouraged in clinical practice to ensure timely clinical intervention.

## Data Availability

All relevant data are within the manuscript and its Supporting Information files.

## Acknowledgments

None

## Availability of data

All studies included in this systematic review and meta-analysis are published and freely available.

